# Theta Oscillatory State-Adaptive Subthalamic Stimulation Modulates Decision-Making under Risk and Uncertainty in Parkinson’s Disease

**DOI:** 10.1101/2025.11.11.25339833

**Authors:** Linbin Wang, Zhitong Zeng, Jingwei Li, Luis Manssuer, Peng Huang, Yixin Pan, Dianyou Li, Valerie Voon

## Abstract

Risk-taking underpins everyday decision-making and is often dysregulated in neuropsychiatric disorders. Theta oscillations in the subthalamic nucleus (STN) have been implicated in conflict, risk evaluation, reward processing and impulsivity, but their causal role in risky choice remains unclear. We aimed to determine whether targeted modulation of STN theta dynamics can influence decision-making under risk and uncertainty. In a randomized, double-blind study of 20 patients with Parkinson’s disease (PD), we applied bilateral subacute STN stimulation at 5 Hz, 130 Hz, or sham during a risk-taking card task. In a separate cohort of 9 perioperative patients, we implemented a closed-loop brain–machine interface to deliver state-dependent theta stimulation. Bilateral subacute STN stimulation at either frequency did not alter risky choice behaviors although 5 Hz stimulation hastened drift rates and 130 Hz stimulation slowed drift rates thus decreasing and increasing caution respectively. Critically, theta state–adaptive high frequency acute STN stimulation reduced risky and uncertain betting without affecting reaction times and was associated with decreased theta power, shorter bursts, and attenuated outcome-related responses. These findings demonstrate that theta state–adaptive STN stimulation causally shapes decision-making under risk and uncertainty, supporting oscillatory closed-loop DBS as a promising neuromodulation strategy for impulsivity and psychiatric disorders.

## Introduction

The evaluation of risk is a fundamental component of human decision-making, weighing potential rewards against possible losses in everyday choices. Risk-taking occurs both under risk, where probabilities are known, and under uncertainty, where probabilities are incomplete or unknown^1^. For example, playing a card game represents a decision under risk because the odds of winning are fixed and known, while investing in a new business venture illustrates a decision under uncertainty, since future outcomes and their probabilities are unknown. Propensity for risky choice represents a facet of decisional impulsivity, as opposed to motor impulsivity, and is thought to engage a distributed cognitive–limbic network involving the ventromedial prefrontal cortex (PFC), dorsal anterior cingulate cortex (ACC), striatum, and anterior insula^2^. Dysregulation of risk-taking contributes to maladaptive choices across neurological and psychiatric conditions, motivating a mechanistic account that can translate to intervention^3,4^.

The subthalamic nucleus (STN) sits at a pivotal inhibitory node within cortical-basal ganglia circuitry and is implicated in heterogeneous forms of impulsivity—including decisional risk taking—across overlapping motor, cognitive and limbic networks^5–7^. Deep brain stimulation (DBS) of the STN is an established therapy for Parkinson’s disease (PD) and obsessive–compulsive disorder (OCD) but can also modulate impulsivity phenotypes and may elicit neuropsychiatric effects^8–10^. The cognitive-limbic STN involvement is particularly associated with increased hypomania characterized by euphoria and risk seeking and impulse control disorders (ICDs), whereas dorsal motor STN stimulation may temper these effects^6,11–13^. Previous DBS studies in PD patients have shown that high-frequency STN-DBS can worsen premature responding in highly uncertain or time-constrained contexts indicative of increased motor impulsivity^14–18^, yet paradoxically produces either no change or a reduction in risky choices when probabilities are explicit or uncertainty is high^19–21^.

STN theta dynamics are implicated in conflict processing^22–24^, response execution^25,26^, reward and risk evaluation^27–31^, and impulsivity^32–34^. Prior studies in PD patients on conflict-related decision-making consistently show increased STN theta activity during decision conflict, associated with an expanded decision boundary and more deliberate choices^15,35,36^. Parallel evidence from economic decision-making tasks indicates that STN theta activity during deliberation relates to cautious strategies and risk avoidance, particularly in PD patients with pathological gambling^37,38^. Converging findings link STN theta to impulsivity and ICDs in PD with peak theta power observed at ventral contacts in line with oscillatory signatures of impulsivity and the behavioral effects of DBS^32,33^. Intracranial and EEG recordings from perioperative PD patients further show that high frequency STN DBS applied in an intermittent time-locked manner during the decision phase can reduce risk-taking under uncertainty, presumably by increasing STN theta activity with faster rates towards the avoid bet boundary^21^. Together these results highlight STN theta dynamics as a latent physiomarker, though it remains unclear whether this oscillation has causal significance for decision-making under risk and uncertainty.

Demonstrating a causal contribution of neural oscillations poses substantial methodological challenges, particularly in human studies. Evidence from recent work indicates that stimulating the subthalamic nucleus (STN) at theta frequencies can perturb theta oscillations and improve conflict resolution and executive functions with increased frontal activation and network connectivity in PD patients^39–41^. These observations suggest that theta-band dynamics may play a causal role in conflict processing. Yet, conventional techniques—whether pharmacological or electrical—lack the specificity required to isolate effects on the oscillation of interest or fail to account for the influence of state dynamic changes. One promising strategy is the adoption of closed-loop brain–machine interfaces (BCI), in which theta activity is monitored in real time and used to control the application of electrical stimulation.

Here, we set out to establish the causal role of STN theta dynamics in decision-making under risk and uncertainty, and to assess whether targeted modulation can modify behavioral outcomes. First, in a randomized, double-blind study of twenty PD patients, we delivered bilateral subacute STN stimulation at theta (5 Hz), high frequency (130 Hz), or sham during a risk-taking card task to assay frequency-specific effects on risky choices. We expected divergent effects consistent with prior findings: high-frequency stimulation would reduce, whereas theta-frequency stimulation would increase, the propensity for risky decision-making. Second, in nine perioperative PD patients, we conducted a proof-of-concept study implementing a closed-loop brain–machine interface in which ongoing left-STN theta activity was monitored in real time to control right-STN stimulation, thereby linking endogenous theta fluctuations to choice under uncertainty and demonstrating the feasibility of targeted, state-dependent neuromodulation.

## Results

### Subacute STN Stimulation (130 Hz, 5 Hz) on Risk Taking

We conducted a blinded, randomized controlled study comparing 130 Hz versus 5 Hz versus sham subacute STN stimulation, to examine the frequency effects on decision-making under risk and uncertainty (**Fig. 1a & 1b**). Twenty patients with PD (3 females, 17 males; mean age 60 ± 9 years; mean disease duration 11 ± 4 years) participated in the subacute stimulation study. The average time since STN DBS was 12 ± 9 months. Detailed demographic and clinical characteristics are provided in **Table S1**.

**Figure 1.**
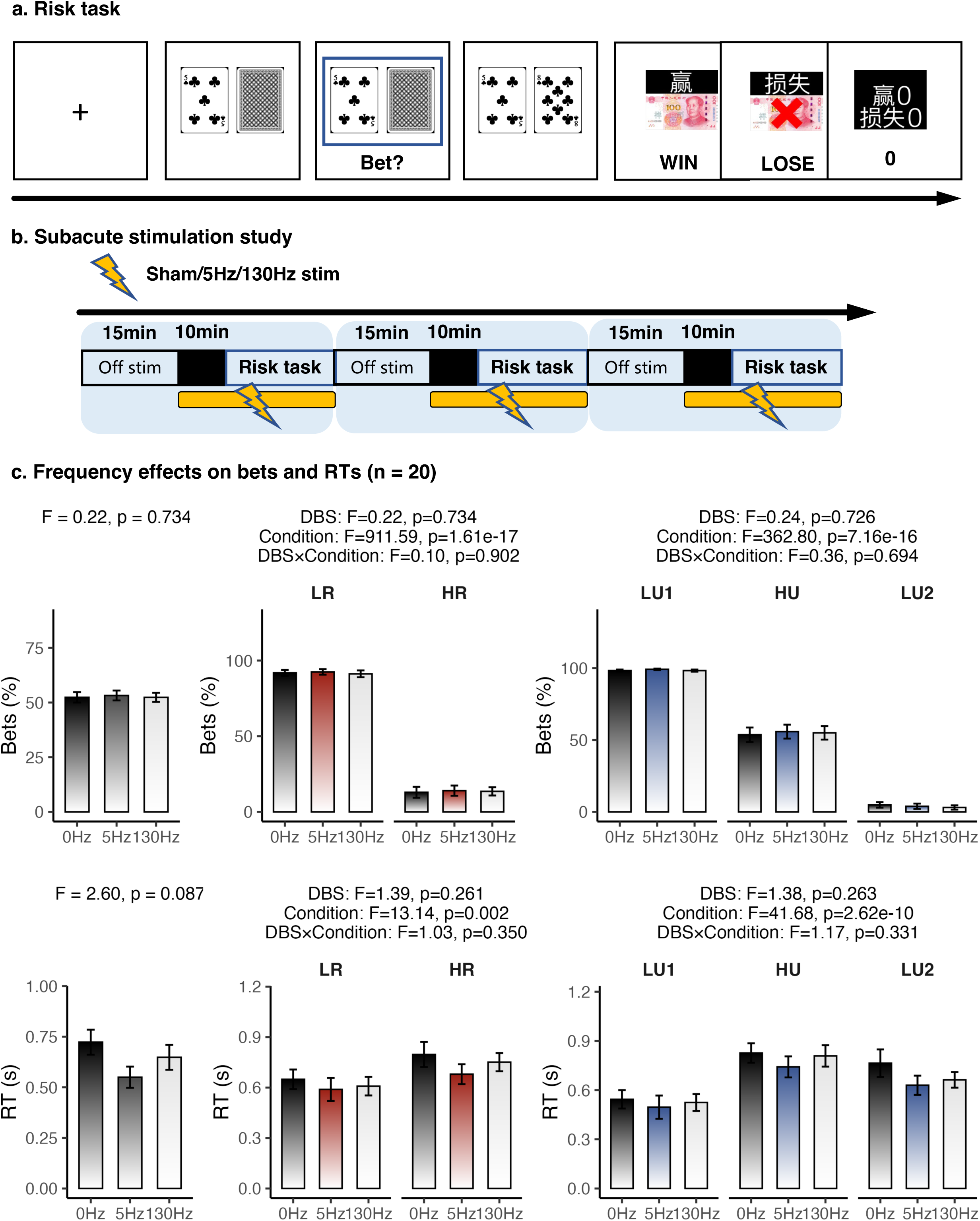
Subacute stimulation study. **a.** Risk task. Participants were asked to decide whether the value of a hidden card exceeded that of a visible card, a design that parametrically manipulated risk (expected payoff) and uncertainty (outcome variability). **b.** Study design. Participants performed the task under three counterbalanced stimulation conditions (5 Hz, 130 Hz, and sham) in a double-blind design. Each condition included wash-in (10min) /wash-out (15min) periods to minimize carryover effects. **c.** Frequency effects on bets and RTs (n = 20). Repeated-measures ANOVA with stimulation frequency (5 Hz, 130 Hz, sham) and condition (high vs. low risk or high vs. low uncertainty) as within-subject factors showed that subacute STN stimulation did not significantly affect betting behavior under either risk or uncertainty. **Abbreviations:** stim, stimulation; min, minutes; RT, reaction time; DBS, deep brain stimulation; LR, low risk; HR, high risk; LU, low uncertainty, HU, high uncertainty.

To explore the frequency effects on bets and reaction times (RTs) under risk and uncertainty, we conducted a repeated-measures ANOVA with stimulation frequency (5 Hz, 130 Hz, sham) and condition (high vs. low risk or high vs. low uncertainty) as within-subject factors.

For bets, there was a robust main effect of risk (F(1,19) = 911.59, p = 1.6e-17) and of uncertainty (F(1,19) = 362.80, p = 7.2e-16). However, stimulation frequency had no significant effect (Risk: F(2,38) = 0.22, p = 0.73; Uncertainty: F(2,38) = 0.24, p =0.73), nor were there significant frequency × condition interactions (Risk: F(2,38) = 0.10, p = 0.9; Uncertainty: F(2,38) = 0.36, p = 0.69) (**Fig. 1c**).

For RTs, we again observed significant main effects of both risk (F(1,19) = 13.14, p = 0.002) and uncertainty (F(1,19) = 41.68, p = 2.6e-10). Yet, as with bets, there were no significant main effects of stimulation frequency (Risk: F(2,38) = 1.39, p = 0.26; Uncertainty: F(2,38) = 1.38, p = 0.26) or interaction effects (Risk: F(2,38) = 1.03, p =0.35; Uncertainty: F(2,38) = 1.17, p = 0.33) (**Fig. 1c**).

We further applied computational modeling using hierarchical drift–diffusion modeling (DDM) analyzing decision boundaries and drift rate separately. We used Bayesian statistics in keeping with DDM outputs where a BF_10_ of 3-10 is indicative of moderate evidence and >10 of strong evidence^42–46^. We revealed significant main effects on drift rate of stimulation frequency (F(2,38) = 9.68, p < 0.001, BF□□ = 23.51) and uncertainty (F(1,19) = 276.45, p < 0.001, BF□□ > 100), as well as a significant interaction between these factors (F(2,38) = 5.74, p = 0.007, BF□□ = 15.97). 5-Hz stimulation increased drift rate overall (slowed information processing towards decision boundaries) with post hoc analyses indicating that 5-Hz STN stimulation significantly increased the drift rate under low-uncertainty conditions (LU: 5 Hz vs. 0 Hz: BF□□ = 5.29; 5 Hz vs. 130 Hz: BF□□ = 215.09; 0 Hz vs. 130 Hz: BF□□ = 0.65; HU: 5 Hz vs. 0 Hz: BF□□ = 0.23; 5 Hz vs. 130 Hz: BF□□ = 0.77; 0 Hz vs. 130 Hz: BF□□ = 0.41) (**Fig. 2a**). For risk, there was a significant main effect of risk (F(1,19) = 324.21, p < 0.001, BF□□ > 100) and a significant risk × stimulation frequency interaction (F(2,38) = 5.42, p = 0.009, BF□□ = 9.18). Post hoc tests revealed that 130-Hz stimulation significantly reduced the drift rate under high-risk conditions (LR: 5 Hz vs. 0 Hz: BF□□ = 0.43; 5 Hz vs. 130 Hz: BF□□ = 0.55; 0 Hz vs. 130 Hz: BF□□ = 0.23; HR: 5 Hz vs. 0 Hz: BF□□ = 0.26; 5 Hz vs. 130 Hz: BF□□ = 4.90; 0 Hz vs. 130 Hz: BF□□ = 4.61) (**Fig. 2b**).

**Figure 2.**
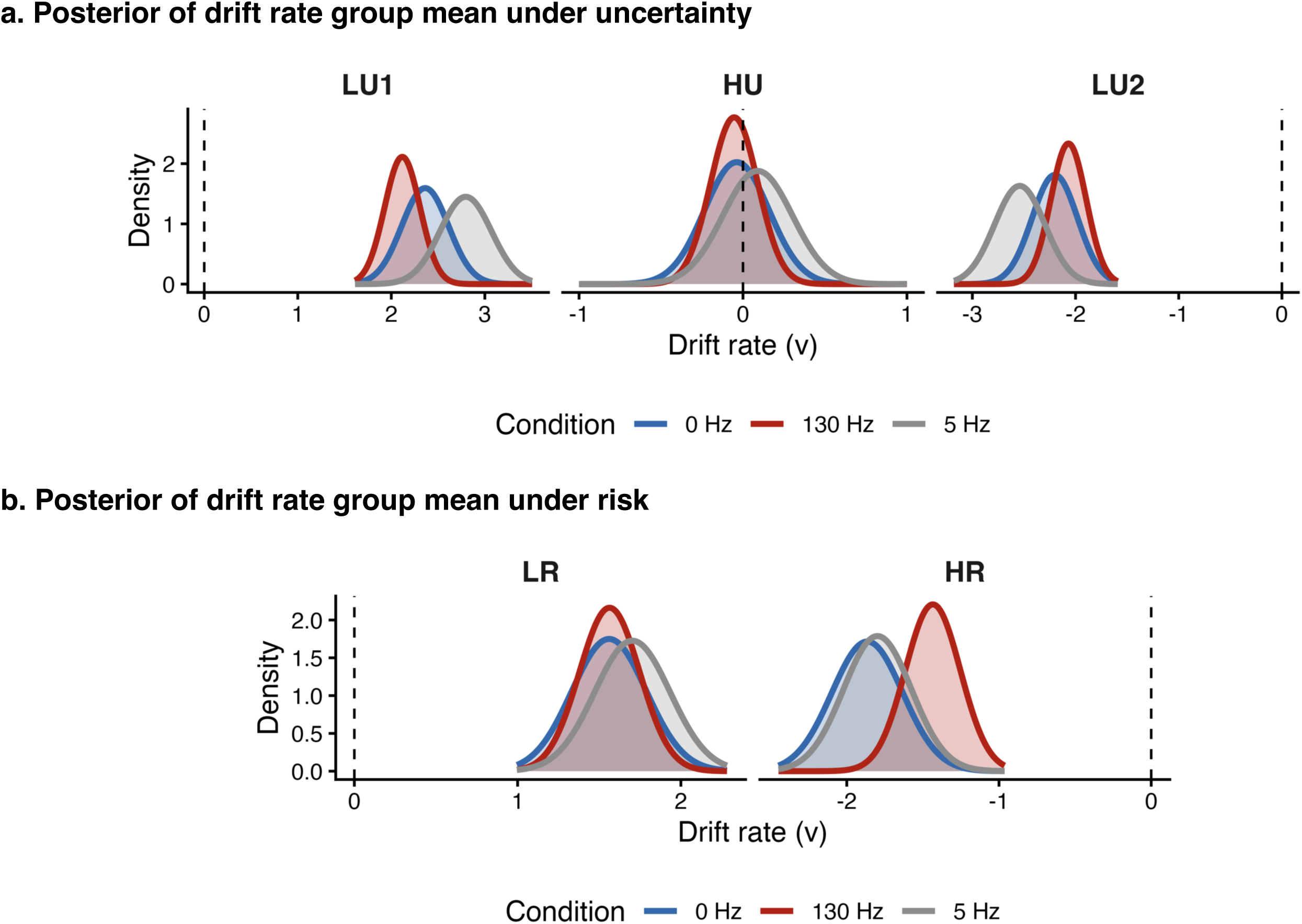
Hierarchical drift–diffusion modeling of decision-making under subacute stimulation. **a.** Posterior of drift rate group mean under high versus low uncertainty (n = 20). Drift rate was significantly influenced by stimulation frequency and uncertainty, with 5-Hz STN stimulation increasing drift rate particularly under low-uncertainty conditions. **b.** Posterior of drift rate group mean under high versus low risk (n = 20). Drift rate was significantly influenced by stimulation frequency and risk, with 130-Hz STN stimulation reducing drift rate specifically under high-risk conditions. **Abbreviations:** LR, low risk; HR, high risk; LU, low uncertainty, HU, high uncertainty.

Together, these findings indicate that subacute STN stimulation did not alter betting behavior under either risk or uncertainty. However, consistent with our hypothesis, subacute 5-Hz STN stimulation globally increased the drift rate index of evidence accumulation towards decision boundaries (hastening decision making) particularly during low uncertainty, whereas 130-Hz stimulation selectively decreased the drift rate (slowing decision making) under high-risk conditions, suggesting latent, frequency-specific modulation of decision making under risk and uncertainty.

### Theta oscillatory state-adaptive Stimulation Decreased Risk Taking

Having shown that subacute STN stimulation at 130 Hz or 5 Hz failed to elicit behavioral effects, we further hypothesized that such effects might be state dependent. To test this, we implemented a closed-loop brain–machine interface in which ongoing left-STN theta activity was monitored in real time to control high frequency right-STN stimulation for one second duration (**Fig. 3a & 3b**). Nine inpatients with Parkinson’s disease performed the risk-taking task during theta oscillatory state-adaptive or sham stimulation (Demographics in **Table S2 & Fig. S1a**).

**Figure 3.**
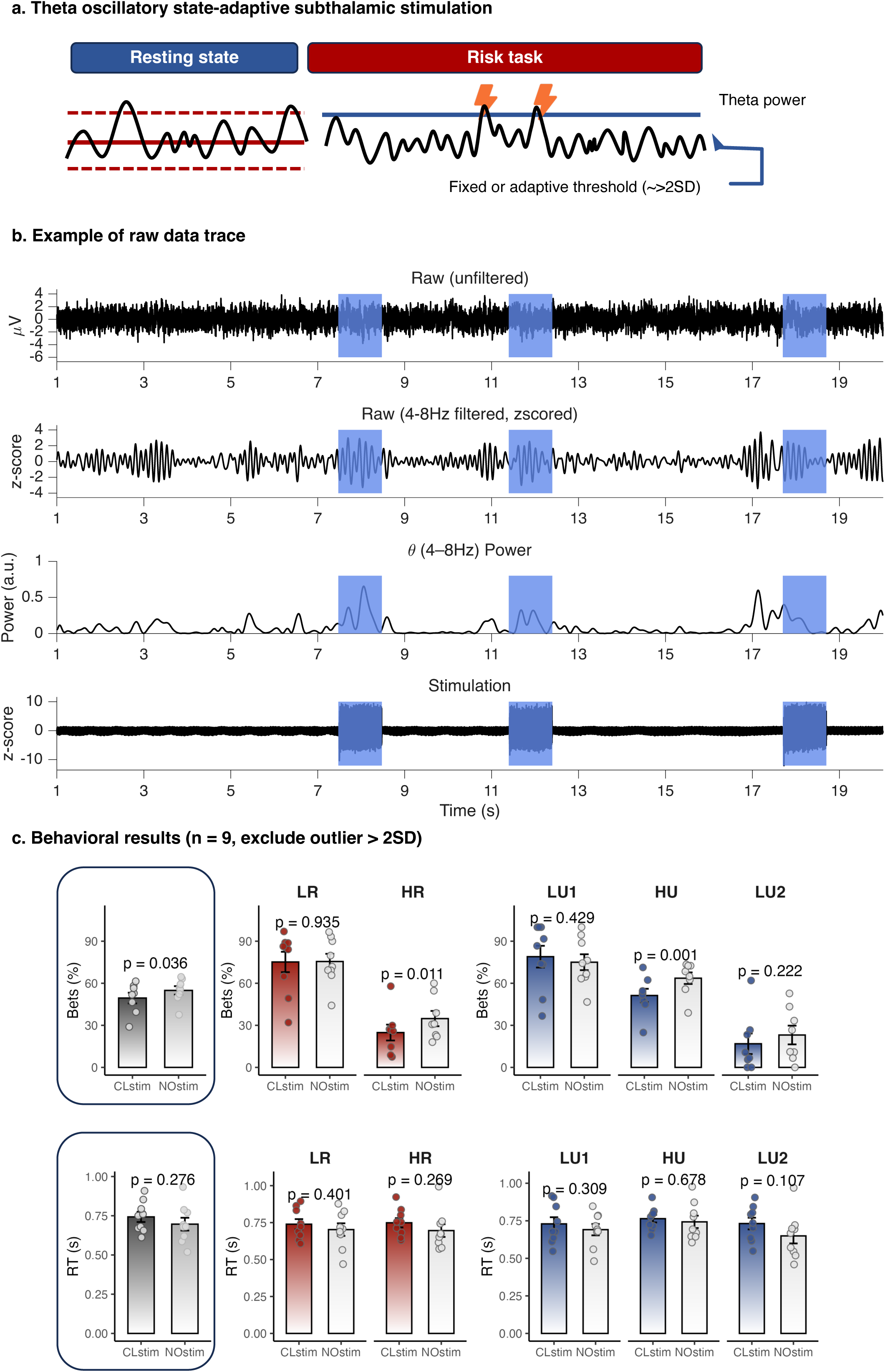
Theta oscillatory state-adaptive subthalamic stimulation and behavioral outcomes. **a.** Illustration of theta oscillatory state-adaptive subthalamic stimulation. Right STN stimulation was delivered at 130 Hz with individualized amplitude settings. STN stimulation was applied at 130 Hz with individualized settings, and adaptive stimulation was triggered in real time based on fixed or adaptive theta-band activity thresholds derived from baseline recordings. **b.** Example of raw data trace with theta oscillatory state-adaptive subthalamic stimulation. From top to bottom: 20-second raw signal; 4–8 Hz filtered signal; Hilbert-transformed theta power; and stimulation artifacts. **c.** Behavioral outcomes (n = 9). Theta oscillatory state-adaptive subthalamic stimulation reduced high-risk and high-uncertainty bets without affecting RTs. p-values from paired tests shown. **Abbreviations:** SD, standard deviation; RT, reaction time; LR, low risk; HR, high risk; LU, low uncertainty, HU, high uncertainty.

As expected, betting behavior scaled with risk, increasing linearly with card value, and varied with uncertainty, peaking at mid-range values and declining at the extremes. Under theta oscillatory state-adaptive STN stimulation, total bets were significantly reduced (t(7) = –2.60, p = 0.036, Cohen’s d = –0.92), as were bets under high uncertainty (t(7) = –5.17, p = 1.29e-3, Cohen’s d = – 1.83) and high risk (t(7) = –3.44, p = 0.012, Cohen’s d = –1.22), relative to sham. By contrast, no significant differences were observed under low risk (t(8) = –0.08, p = 0.94, Cohen’s d = –0.03) or low uncertainty (Low uncertainty 1: t(8) = 0.83, p = 0.43, Cohen’s d = 0.28; Low uncertainty 2: t(7) = –1.34, p = 0.22, Cohen’s d = –0.47). Reaction times did not differ significantly between closed-loop and sham stimulation (all p > 0.05) (**Fig. 3c**).

Together, these findings demonstrate that theta oscillatory state-adaptive STN stimulation selectively reduces risky and uncertain betting behavior without affecting response speed.

### Stimulation Pattern of Theta Oscillatory State-Adaptive Stimulation

Risk taking involves subjective risk evaluation and conflict resolution during the deliberation phase, response execution during the response phase, prediction error monitoring and value processing during the outcome phase. Using theta oscillatory state-adaptive DBS allowed us to determine in which cognitive process stimulation might have an effect. In this paradigm, stimulation was delivered contingent on STN theta power, such that DBS was switched on for one second during periods of high theta power and off during periods of low theta power (**Fig. S2a**).

Notably, theta oscillatory state-adaptive DBS (n = 7) induced a theta-frequency phase reset, reflected by increased cross-trial phase coherence during stimulation relative to pre- (z = -2.20, p = 0.047, rbc = -0.929) and post-stimulation (z = 2.20, p = 0.047, rbc = 0.929) periods, while no difference was found between pre- and post- stimulation periods (z = -1.01, p = 0.375, rbc = - 0.429) (**Fig. S2b**).

Temporal variability and state-dependent roles of theta power resulted in DBS being delivered at different time windows across trials. We then assessed the dynamics of stimulation delivery over trials. A dynamic stimulation analysis (200ms) aligned to the decision, response, and reward outcome respectively revealed that stimulation delivery peaked (∼5% of all trials) peri-response, approximately 500ms before card outcome presentation (**Fig. S1b**).

### Stimulating at the Decision Phase

Our prior work demonstrated that intermittent, decision-phase–locked high-frequency STN DBS enhanced cue-elicited theta activity, driving faster trajectories toward the avoidance boundary and reducing risk-taking under uncertainty in PD patients^21^. Accordingly, we posited that theta oscillatory state-adaptive DBS at decision phase would enhance STN theta activity and shift betting behavior under risk and uncertainty. To test this hypothesis, cue-elicited STN theta power during the decision phase was compared between closed-loop DBS and sham at both subject and trial levels.

Given trial-to-trial jitter in stimulation delivery, we evaluated theta power across the 3-s decision phase to minimize temporal variability. At the subject level (n = 6), cue-elicited theta power showed a marginal overall decrease (t(5) = –1.78, p = 0.094, rbc = –1), reaching significance during high-uncertainty (t(5) = –2.20, p = 0.031, rbc = –1) and low-risk trials (t(5) = –2.20, p = 0.031, rbc = –1), but not during low-uncertainty or high-risk trials (both p > 0.05) (**Fig. 4a**). At the trial level, total theta power was computed over the 3-s decision phase for each trial and normalized within subjects to enable cross-subject comparison. For comparison, we also examined beta power, as STN beta activity is known to be modulated by continuous 130Hz DBS in PD patients. Theta oscillatory state-adaptive DBS significantly reduced theta power relative to sham (t(1704) = –3.113, p = 0.002, Cohen’s d = -0.151) (**Fig. 4a**), whereas beta power remained unchanged during the decision phase at both the subject level (all p > 0.05 across high/low risk and uncertainty conditions) and the trial level (t(1704.0) = –1.44, p = 0.149, Cohen’s d = –0.07) (**Fig. S3a**). On an exploratory basis, we examined whether decision-phase theta power correlated with betting behavior by pooling closed-loop DBS and sham stimulation datasets (n = 12). Correlation analysis revealed that theta power during decision phase were positively associated with betting under uncertainty (Spearman correlation: r = 0.587, p = 0.049, Fisher’s z = 0.674; Pearson correlation: r = 0.577, p = 0.05, Fisher’s z = 0.658). These findings demonstrate that theta oscillatory state-adaptive DBS reduces theta power during the decision phase, which is associated with decreased betting under uncertainty.

**Figure 4.**
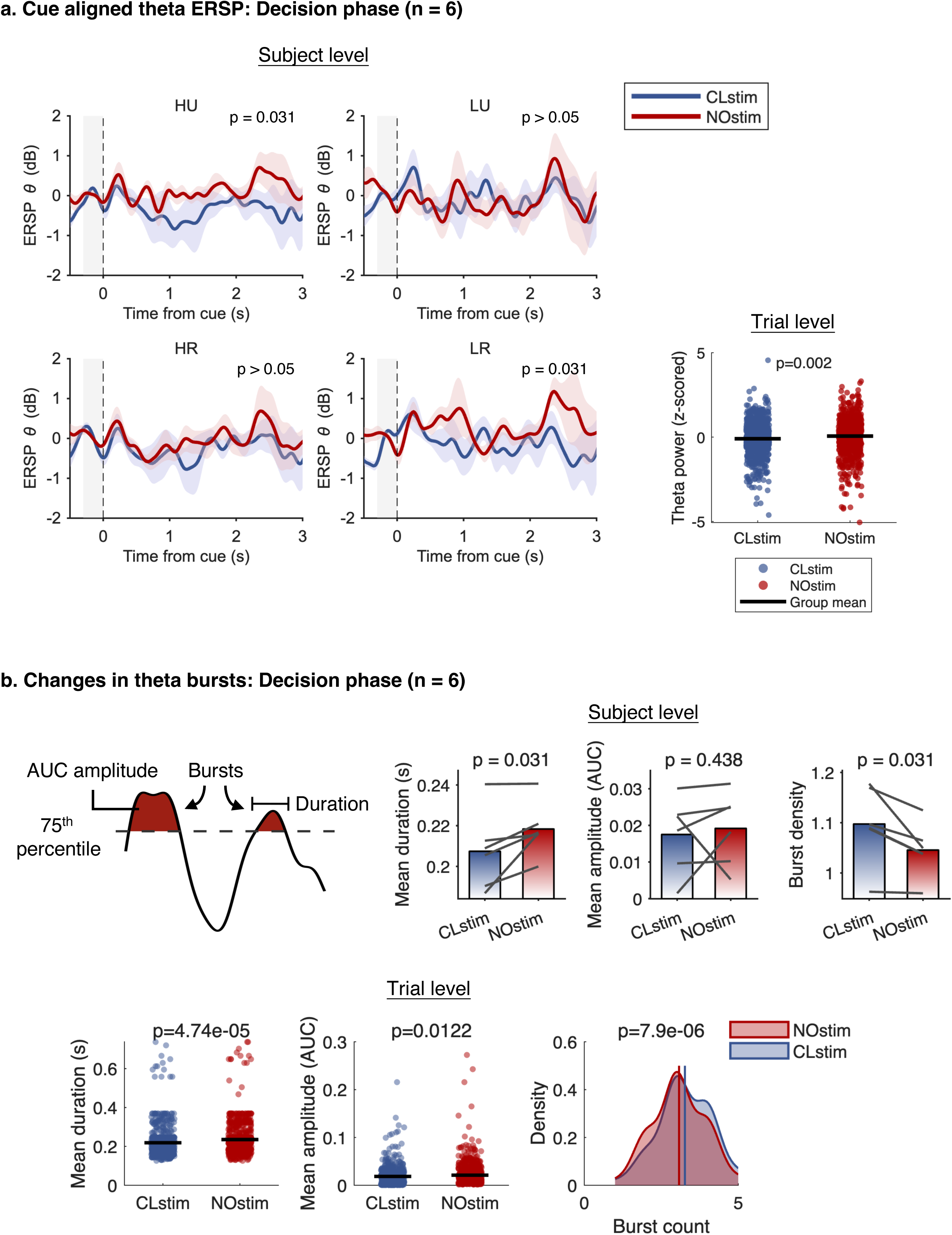
Theta dynamics during decision phase under theta oscillatory state-adaptive subthalamic stimulation. **a.** Left: Cue-aligned theta ERSP for high vs. low uncertainty and risk conditions during the decision phase (n = 6, subject level), showing reduced theta-band STN activity under theta oscillatory state-adaptive subthalamic stimulation compared to sham. Paired-test p-values across the 3-s decision phase are indicated on the plot. Right: Cue-aligned theta ERSP during the decision phase (n = 6, trial level), showed reduced theta-band STN activity under theta oscillatory state-adaptive subthalamic stimulation compared to sham. **b.** Theta burst metrics (AUC, amplitude, and duration) during the decision phase (n = 6), demonstrating reduced burst duration and increased burst density under theta oscillatory state-adaptive stimulation relative to sham (upper: subject level; lower: trial level). Burst detection threshold set at the 75th percentile. **Abbreviations:** ERSP, event-related spectral perturbation; Nostim, no stimulation; CLstim, close loop stimulation; AUC, Area Under the Curve; LR, low risk; HR, high risk; LU, low uncertainty, HU, high uncertainty.

Beta-triggered closed-loop DBS has been shown to truncate long beta bursts, shifting activity toward shorter (higher density), lower-amplitude bursts, with burst duration correlating with motor impairment^47^. By analogy, our theta oscillatory state-adaptive DBS system tracks theta bursts to guide stimulation, leading us to hypothesize that it may similarly modulate theta burst duration, amplitude, and density during the decision phase, thereby shaping betting behavior.

Theta bursts were defined analogously to beta bursts, using the 75th percentile of the amplitude distribution as threshold^48^. At the subject level, we found that theta oscillatory state-adaptive DBS significantly increased theta burst density (t(5) = 2.201, p = 0.031, rbc = 1) while reducing burst duration (t(5) = –2.201, p = 0.031, rbc = -1), with amplitude remaining unchanged (t(5) = –0.943, p = 0.438, rbc = –0.429). In contrast, beta burst density, duration, and amplitude were unaffected (all p > 0.05) (**Fig. 4b & Fig. S3b**). At the trial level, theta bursts were extracted for each trial and compared between closed-loop DBS and sham stimulation. Consistent with the subject-level results, theta oscillatory state-adaptive DBS significantly reduced burst duration (t(1535) = -4.070, p = 4.93e-05, Cohen’s d = -0.208) and amplitude (t(1535) = -2.493, p = 0.013, Cohen’s d = -0.128) while increasing burst density (t(1535) = 4.485, p = 7.82e-06, Cohen’s d = 0.229) (**Fig. 4b**).

Together, we found that theta oscillatory state-adaptive DBS specifically reduced theta power and burst duration while increasing burst density during the decision phase. Thus, theta bursts were less sustained and intense and more fractured. Lower theta power during decision phase were associated with lower betting under uncertainty.

### Stimulating at the Outcome Phase

Having said that stimulation delivery peaked ∼500ms before the outcome phase, theta oscillatory state-adaptive DBS may also alter betting behavior by influencing the outcome phase. Previous studies have implicated increased STN theta activity in value processing (reward/loss)^21,27,28,30,31^. We therefore hypothesized that theta oscillatory state-adaptive DBS would modulate STN theta activity during reward and loss processing. To test this, we compared outcome-elicited STN theta activity between closed-loop DBS and sham stimulation during bet (reward/loss) and no-bet trials.

We observed an increase in theta activity when comparing bet (reward/loss) versus no-bet trials at both the subject (200-700ms, all uncorrected p < 0.05, sliding Wilcoxon signed-rank test) and trial (100–400ms, all uncorrected p < 0.05, sliding student t test) levels, replicating our previous findings^21,49^. At the subject level, no significant differences were found between closed-loop DBS and sham stimulation (p > 0.05), although a relative reduction in theta activity during closed-loop DBS was observed. At the trial level, a cluster-based permutation test showed that closed-loop DBS significantly reduced outcome-elicited theta activity (50–450ms, corrected p = 0.032). In contrast, no significant changes were observed during no-bet trials, either between closed-loop and sham DBS or between the pre-outcome and post-outcome phases (all p > 0.05) (**Fig. 5**).

**Figure 5.**
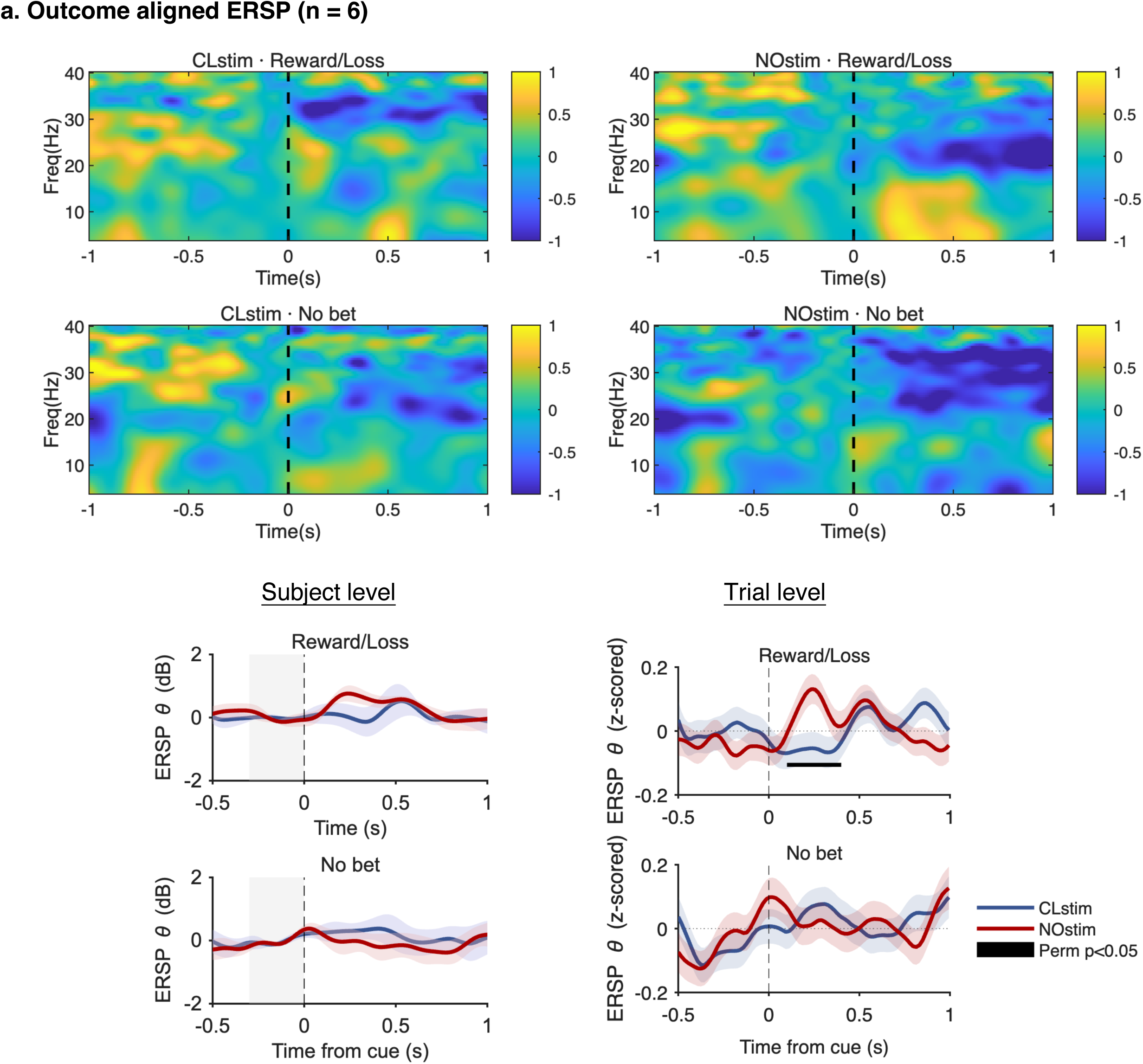
Theta dynamics during outcome phase under theta oscillatory state-adaptive subthalamic stimulation. Upper: Time–frequency plots of outcome-aligned activity under reward/loss and no-bet conditions during sham and theta oscillatory state-adaptive STN stimulation. **Lower:** Outcome-aligned theta-band ERSP during outcome phase (n = 6), shown at the subject level (left) and trial level (right), comparing theta oscillatory state-adaptive stimulation to sham. Horizontal black lines indicate significant clusters identified using a sliding Wilcoxon signed-rank test (200-ms window, 100-ms step) with cluster-based permutation correction. **Abbreviations:** ERSP, event-related spectral perturbation; Nostim, no stimulation; CLstim, close loop stimulation.

Together, these results suggest that theta oscillatory state-adaptive DBS attenuates the early STN theta response to reward and loss during bet trials, but has no effect on no-bet trials.

## Discussion

To summarize, subacute open-loop stimulation at clinical frequencies (130 Hz) slowed drift rates in high-risk conditions thus increasing cautious behavior and at theta (5 Hz) frequency hastened drift rates in low uncertainty conditions thus decreasing cautious behavior. Thus, although subacute stimulation did not alter risk taking behavior, drift diffusion models highlight a dissociable effect of stimulation frequency on risky decision-making behaviors consistent with our hypotheses.

State-dependent theta oscillatory state-adaptive DBS at high frequency specifically modulated STN theta dynamics, leading to decreased risky and uncertain betting behavior without affecting response speed. We reveal parallel mechanisms of effect: (i) reduced theta power and burst duration with increased burst density during the decision phase, whereby lower theta power correlated with reduced betting behavior; and (ii) attenuation of outcome-elicited theta responses during reward/loss processing. The findings demonstrate that adaptive, oscillatory state-dependent modulation of STN theta activity can shape decision-making under risk and uncertainty, positioning theta oscillatory state-adaptive DBS as a promising framework for next-generation neuromodulation strategies targeting pathological cognitive processes in the human brain.

### Effects of Subacute STN Stimulation

In a randomized, double-blind design, we demonstrate that subacute STN DBS at either conventional (130Hz) or theta (5Hz) frequencies does not alter risk-taking behavior. However, using computational HDDM, subacute 5Hz STN stimulation increased the drift rate under conditions of high certainty, hastening decisions towards the risk-taking boundary with lower cards and hastening decisions towards the safe boundary with higher cards. Subacute 130Hz stimulation selectively decreased the drift rate under high-risk conditions, slowing decisions towards the safe boundary. Thus, although we did not show a change in risk taking behavioral choices, our latent computational outcomes are consistent with our *a priori* hypotheses. Theta frequency stimulation appears to decrease caution, hastening decision tendencies during conditions of high certainty whereas high frequency stimulation increases caution, slowing decision towards the safe boundary in risky choices. These findings contrast with our previous findings showing that intermittent, time-locked acute one-second high frequency STN stimulation during the decision phase reduces risk-taking under uncertainty, as well as with other small pilot studies with chronic high frequency stimulation^19,21,38^. Acute one second stimulation specifically timed and causal to the decision phase may differ from subacute stimulation lasting 15 minutes as per our study and from chronic stimulation. Our subacute study design may be of insufficient duration to have a pronounced effect on risk taking behaviors relative to chronic cumulative stimulation and potentially also influence multiple cognitive processes during the task, thus influencing only latent variables. Theta frequency stimulation appears to enhance caution across decision types towards both the risky and safe boundaries suggesting a broader more generic effect on decision tendencies whereas high frequency stimulation shifts caution specifically towards the safer boundary suggesting a more specific effect on risk taking. Further studies to examine the effect on physiology are indicated as the effects of subacute theta stimulation and high frequency stimulation, while dissociable are not in the opposite direction.

### Oscillatory State-Adaptive Dependent Neuromodulation for Risk Taking

The subacute findings underscore that stimulation effects may be best captured in a state-dependent rather than chronic manner. Adaptive closed-loop DBS has advanced treatment of PD motor symptoms by enhancing efficacy and reducing side effects, using STN beta and more recently gamma activity as control signals^50–53^. Extending this approach to cognition is critical, as cognitive interventions are double-edged and unintended interference with non-targeted processes could be detrimental in human patients. We have previously implemented a cognitive state-dependent stimulation paradigm that showed promise in reducing risk-taking in a controlled laboratory setting^21^. However, real-world application of this approach is challenging, as determining cognitive state in real time remains difficult. In this context, identifying robust biomarkers, developing real-time decoding methods for cognitive state, and using these biomarkers to drive neuromodulation may provide a viable alternative. In this proof-of-concept study, we show that theta oscillatory state–adaptive STN stimulation reduces risk-taking under both uncertainty and risk highlighting its potential as a targeted neuromodulation strategy for cognitive control.

There are several plausible mechanistic explanations underlying our findings explained in the following. These can be separated into the effects of theta-adaptive DBS on cognitive-physiological mechanisms on risk taking including an influence of inhibitory processes, reversal of the relationship between physiology and decision making and an impact on cost-benefit and reinforcement learning processes. First, an influence of inhibitory processes may be relevant to risk taking. Our in-patient implementation of theta state–adaptive STN stimulation showed stimulation was broadly triggered across the risk-taking process, peaking near response. This converges with evidence linking STN theta to response execution and lowered decision thresholds. Our findings may thus implicate response inhibition processes, one of the most critical functions of the STN and potentially intrinsically relevant to risk taking and reinforcing theta as a robust causal biomarker of this process^22–26,54^. We had previously shown that acute stimulation of the ventral associative-limbic STN along with pre-supplementary motor area and STN white matter tracts is more closely linked with risk taking whereas the dorsal motor STN and supplementary motor area and STN tracts are associated with risk aversion.

Thus, the pre-supplementary motor area – STN network implicating both response inhibition and motor preparation networks appears to be associated with differential risk-taking tendencies with stimulation relative to the supplementary motor area – STN network implicated only in motor preparation. We had previously suggested that acute STN stimulation influencing this response inhibition pre-supplementary motor area network may have an influence on risk taking by putting on ‘brakes’ and slowing decision processes to carefully evaluate the likelihood of a reward or punishment^21^. Notably, our use of fixed or adaptive thresholds for theta power inherently tracks the highest theta peaks and may inadvertently stimulate during non-relevant peaks, underscoring the need for personalized decoding tailored to the specific cognitive process of interest in future studies.

Intriguingly, we found that theta state–adaptive STN stimulation reduced theta power, with lower power during decision phase linked to reduced betting under uncertainty. This contrasts with our prior finding that intermittent time-locked stimulation increased STN theta correlating with decreasing bet rates and accelerated drift rate toward the avoid-bet boundary^21^. Notably, in our original study, time-locked stimulation was applied during the first second of the decision phase, preceding the rise in theta, whereas our state–adaptive stimulation in this study was delivered once theta had risen, suggesting distinct mechanisms from both intermittent and adaptive stimulation. In conflict processing, STN-DBS reverses the correlation between medial prefrontal EEG theta activity and decision threshold, such that higher prefrontal theta is associated with reduced evidence accumulation, reflecting disrupted mPFC–STN communication under continuous STN stimulation ^15^. Theta state–adaptive STN stimulation may have a similar effect, selectively inhibiting STN theta power and reversing this relationship with risk taking choices and presumably reduces evidence accumulation with more cautious decision-making towards risk aversion. This is relevant to PD patients with impulse control disorders (ICDs), who exhibit abnormally elevated resting-state STN theta activity worsened by conventional STN-DBS and levodopa treatment, whereas theta state–adaptive high-frequency STN stimulation may offer an alternative^32,33^.

Another account for this observation is theta state–adaptive high-frequency stimulation may disrupt physiologically meaningful theta bursts during decision making, thereby interfering with normal prefrontal–STN communication. Theta burst analysis, in parallel with beta bursts, shows that theta state–adaptive high-frequency stimulation disrupts long theta bursts into shorter, lower-amplitude bursts. Ensuring a sufficiently long and complete theta ramp may be critically important; disruption of these bursts could lead to either a sudden collapse of the decision boundary or impaired response inhibition, leading to irrational and near-random choice behavior^51^.

Finally, risk-taking particularly under uncertainty involves a cost–benefit evaluation, and learning from reward or loss outcomes may shape the decision-making boundary. Low-frequency oscillations in the STN have been associated with reward anticipation, reward magnitude^55^, and effort for reward^30,31^, with theta activity enhanced across both win and loss outcomes irrespective of monetary value^21,27^. Thus, decreases in theta power during the decision phase with theta-adaptive DBS might selectively influence the anticipation of reward, thus shifting towards a greater cost-benefit decision bias and enhancing risk aversion. We further observed that theta activity to both reward and loss outcomes was enhanced following sham stimulation but not following theta state–adaptive high-frequency STN stimulation. These findings are consistent with previous work and highlight diminished outcome sensitivity and perhaps decreased outcome learning possibly shifting motivated reinforcement learning from feedback toward a random choice policy (i.e. closer to 50%), which may partly explain the observed reduction in betting^21,31^. Thus, alternate explanations in our risk aversion findings might lie in a shift towards a greater cost-benefit anticipatory bias or in decreased learning from feedback.

### Implications and Challenges for Closed-Loop DBS in Daily Decision-Making

This work is particularly relevant to recent advances in adaptive DBS and the rise of BCI, challenging the field to explore how state-dependent, closed-loop neuromodulation can be leveraged to benefit a broader population with psychiatric disorders marked by pathological cognitive processes, extending its application beyond motor symptom management.

While this study provides a valuable proof of concept, key limitations need to be addressed to bridge the gap toward real-world implementation. As mentioned above, STN theta dynamics are implicated in multiple stages of risk-taking, including conflict processing, response execution, reward and risk evaluation and learning from feedback. It will be crucial to identify and dissociate these theta biomarkers—potentially by integrating additional neural signals or connectivity measures—and determine which are most causally relevant for modulating risk-taking behavior^56–58^. In this context, advanced decoding algorithms are needed to accurately track relevant behavioral states and disentangle risk-related neural biomarkers from unrelated cognitive processes. While our closed-loop results are promising, they underscore the challenge of accurately estimating oscillatory state in real time, as the Hilbert transform requires large data windows around the time of interest and is susceptible to edge effects. Practical implementation will require the development of fast, stable algorithms that minimize decoding delays and deliver stimulation with sufficient temporal precision to modulate the targeted process. Furthermore, although this study implemented an adaptive threshold to personalize the theta biomarker that is potentially better suited for real-world application, future studies may consider incorporating a deeper level of precision through individualized calibration and artificial intelligence (AI)–based biomarker identification, which may further enhance outcomes.

### Limitations

The study is not without limitations. First, although current was matched across frequencies in the subacute study, 5Hz stimulation delivered substantially less total energy than 130Hz, likely leading to an underestimation of its effects. Note that the total electrical energy delivered (TEED) cannot be equalized without altering the volume of tissue activated (VTA). Optimal washout intervals and stimulation durations remain undefined: prior studies with cognitive studies at 5Hz report washouts up to 15 minutes^39–41,59^. Systematic investigation of stimulation dose and timing will be essential to accurately interpret frequency-specific effects on risk-taking and impulsivity. Second, the theta oscillatory state–adaptive DBS study had a very small sample size, which may have increased the risk of Type I error and limited the robustness of the findings. However, we emphasize the rarity of patients who have undergone externalization and further emphasize that our sample size is consistent with other studies on closed loop stimulation and further that our behavioral data all consistently show the same direction of effect^21,60,62,64^. Furthermore, the study lacked an ideal control with random stimulation, leaving open the possibility that the observed effects were due to stimulation itself rather than the theta state–adaptive system. Nevertheless, this alternative explanation appears less likely given that no behavioral changes were observed with subacute continuous 130Hz or 5Hz stimulation.

## Conclusion

In conclusion, our study highlights the potential for subacute theta and high frequency stimulation and adaptive, theta state–dependent modulation of STN activity in shaping decision-making under risk and uncertainty, supporting theta oscillatory state–adaptive DBS as a promising strategy for next-generation neuromodulation targeting psychiatric disorders.

## Materials and Methods

### Participant recruitment and criteria

#### Study I

This was a prospective double-blinded randomized study designed to compare the subacute effects of bilateral 5Hz versus 130Hz versus sham deep brain stimulation (DBS) to the subthalamic nucleus (STN) on decision making under risk and uncertainty in Parkinson’s disease (PD) patients. The study was conducted within the Department of Neurosurgery, Center for Functional Neurosurgery, Ruijin Hospital, Shanghai Jiao Tong University School of Medicine, from June to December 2022. Twenty patients with advanced PD who have undergone bilateral STN DBS for more than 6 months were enrolled in the study. These patients met the criteria: (1) age > 18 years old; (2) cognitive intact (mini-mental state examination score greater than 24); (3) at least 6 months after DBS surgery; (4) Lead-DBS-based electrode reconstruction confirmed at least one contact located within each STN in both hemispheres.

#### Study II

Nine inpatients who had undergone STN DBS surgery at Ruijin Hospital, Shanghai Jiao Tong University School of Medicine, were recruited between July 2024 and July 2025. Inclusion criteria for the study were: (1) age 18–75 years, right-handedness, (2) idiopathic Parkinson’s disease, Hoehn–Yahr stage 2–4 in the off-medication state, (3) Mini-Mental State Examination (MMSE) score >24 or Montreal Cognitive Assessment (MoCA) score ≥18, (4) Perioperative Lead-DBS-based electrode reconstruction confirmed at least one contact located within each STN in both hemispheres and (5) normal vision and hearing. Exclusion criteria included (1) major psychiatric disorders (e.g., schizophrenia, bipolar disorder) meeting DSM-5 criteria; (2) other major neurological disease; (3) unstable vital signs; (4) post-operative complications detected via MRI; or (5) post-operative conditions (e.g., delirium) potentially interfering with task performance.

The studies were approved by the Ethics Committee of Ruijin Hospital, Shanghai Jiao Tong University School of Medicine, and all participants provided written informed consent according to the Declaration of Helsinki. Gender and ethics were self-reported by patients.

### Experimental Task: Risk-Taking Task

Value-based decision-making under risk and uncertainty was assessed using a computerized card task. The task was programmed in MATLAB R2018a with Psychtoolbox 3.0 and presented on a 23.8-inch monitor (1,920 × 1,080 resolution; refresh rate: 75Hz). Participants were seated ∼75 cm from the screen.

On each trial, participants were presented with two virtual card decks (card values 1-10) displayed side-by-side at the center of the screen with one face-up and one face-down. Participants had to bet whether the hidden deck was higher or lower than the visible deck. The design exploited the fact that risk (expected loss-reward slope) increases linearly with card number, while uncertainty (outcome variance) peaks for middle values and is minimal at the extremes.

Before starting the main task, participants received detailed instructions and completed 10 practice trials to minimize practice effects. Each trial began with a fixation cross (1–1.5 s inter-trial intervals), followed by the two card decks. Participants were instructed to respond quickly and try to win as much as possible. Responses were made within 2 s by pressing one of two buttons (right thumb = “higher”; left thumb = “abstain”). Following each response, 1-second arresting feedback was provided: correct guesses earned 1-yuan, incorrect guesses lost 1-yuan, and abstentions resulted in no gain or loss.

In Study I (behavioral version), participants responded immediately upon deck presentation, with the hidden card revealed directly afterward. There were 50 trials total, with 5 trials each for card values 1-10.

In Study II (BCI version), the task was adapted to dissociate cognitive processes for electrophysiological analysis and to extend the decision phase. Following fixation, decks were displayed for 3.1 seconds, after which a response cue (a box around the decks) prompted participants to respond within 2 seconds. This separated decision process from response process. A 0.7-s delay was introduced after the response to allow motor-related activity to dissipate, followed by a 1-s delay after the hidden card was revealed to present feedback, thereby dissociating prediction error monitoring from reward/loss processing. In P1 and P2, the task included 130 trials (13 per card value). In subsequent patients, uncertain trials (card values 4–7) were increased, yielding 150 trials: 30 each for card values 4–7, and 5 each for values 1–3 and 8–10. This adjustment aimed to better capture decision-making under uncertainty and to reduce the influence of obvious outcomes that are less responsive to taDBS.

Local field potentials (LFPs) were recorded using a BrainAmp amplifier (Brain Products), with event markers sent from MATLAB to Recorder software via a parallel port, enabling precise synchronization of behavioral and neural data.

### Study I: Subacute Stimulation

#### Study Design

After recruitment, patients underwent comprehensive assessments under three bilateral stimulation conditions (5 Hz, 130 Hz, and sham stimulation) while in their regular on-medication state. Both assessors and patients were blinded to the stimulation condition. Each condition was separated by a 15-minute off-stimulation interval to minimize post-STN DBS carryover effects. Stimulation frequency order was pseudo-randomized and counterbalanced across all six possible sequences. The risk-taking task commenced 10 minutes after each frequency change to allow for both stimulation wash-in and wash-out, with stimulation remaining on during task.

#### Stimulation Configuration

Subacute stimulation was delivered in a monopolar configuration, with the selected lead contact (cathode) located within the STN and the implanted pulse generator (IPG) serving as the anode. Contact placement was verified using fused preoperative MRI and postoperative CT images in the Lead-DBS v2 toolbox (https://www.lead-dbs.org)^61^. This contact was typically positioned more ventrally than the active contact used clinically; therefore, the motor symptom assessments under stimulation did not reflect patients’ clinical therapeutic status.

Stimulation parameters matched the clinical voltage setting with a fixed pulse width of 90 μs. Notably, the total electrical energy delivered (TEED) for 130 Hz stimulation was 26 times greater than that for 5 Hz. We matched voltage instead of TEED in this study, as matching 130 Hz and 5 Hz markedly altered the volume of tissue activated (VTA) and recruits different brain regions.

#### Clinical Evaluation

Experienced neurologists performed clinical evaluations. Before the experiment, depression and impulsivity were assessed with the use of the Beck Depression Inventory, Second Edition (BDI-II)^63^, and UPPS-P Impulsive Behavior Scale (UPPS-P)^67^. Higher scores in BDI-II indicated more severe depression. The UPPS-P is a 59-item scale measuring 5 separate impulsivity related traits: positive urgency, negative urgency, sensation seeking, lack of premeditation, and lack of perseverance. Part III of Movement Disorder Society-Unified Parkinson’s Disease Rating Scale (MDS UPDRS-III) for motor symptom severity was assessed repeatedly under three stimulation conditions^65^.

### Study II: Theta Oscillatory State-Adaptive STN Stimulation

#### Surgical and experimental procedure

Bilateral DBS leads were implanted under general anesthesia using MRI-guided targeting (3T, General Electric). Two patients (P1&P2) received Medtronic model 3387 leads (four 1Cmm contacts, 1.5Cmm spacing), and the remaining patients (P3–P9) received SceneRay model SR1202-S leads (eight 1Cmm contacts, 0.5Cmm spacing). MRI scans were acquired for each participant and co-registered with computed tomography (CT) images (General Electric) using the Leksell stereotactic frame to determine target coordinates. During the perioperative period following surgery while the leads were externalized (1–2Cdays after lead implantation), participants underwent intracranial signal recordings synchronized with behavioral assessments and stimulations in the ON-medication state (a minimum of 30Cminutes after their usual dose of dopaminergic medications). They completed risk-taking tasks twice: once without stimulation and once with theta oscillation–dependent adaptive STN stimulation (130Hz) on a separate day.

#### Signal Acquisition

Signals were acquired using a BrainAmp MR amplifier (Brain Products, Germany). This system is compatible with BCI2000 (https://www.bci2000.org/) via the FieldTripBuffer interface, featuring real-time data streaming and processing^66^. Local field potential (LFP) signals were simultaneously recorded at a sampling rate of 500 Hz. A 50 Hz notch filter was applied online to suppress power line interference. The data was reference online using a left mastoid electrode. The ground was placed at the apex nasi.

#### Stimulation

Stimulation was applied unilaterally to the right STN via a SceneRay Model 1510 external pulse generator (Suzhou, China), which had received approval from China’s National Medical Products Administration. The stimulation had a fixed frequency of 130 Hz with biphasic pulse width set at 60 μs. We determined the contact pair for bipolar stimulation using lead-DBS–based lead reconstruction (P1–P2: contact 0 as cathode and contact 1 as anode in bipolar configuration; contact 0 is ventral and contact 3 is dorsal of the four contacts; P3–P9: contact pair 2–3 or 5–4 as cathode–anode in bipolar configuration; contact 0 is ventral and contact 7 is dorsal of the eight contacts) to ensure on-target stimulation. We individualized the stimulation intensity for each participant following a standard protocol described in previous reports. Briefly, we increased stimulation intensity by 1V increments at 130 Hz until paresthesia occurred, then reducing it in 1 mA steps until no side effects remained to maintain blinding. If no paresthesia was reported, we applied a maximum amplitude of 3 mA to avoid VTA spread. We confirmed that participants could not detect the onset, offset, or presence/absence of stimulation.

#### Feature Detection, Threshold and Online Processing Pipeline

The sensing contact pair was determined based on lead-DBS–based lead reconstruction. We selected the first sensing contact closest to the DISTAL atlas-defined STN within 0.5mm, with the second closest contact as the reference.

Stimulation was triggered by detecting peaks in theta-band (4–8 Hz) power in the left STN, as these oscillations are the most consistently reported biomarker of conflict-related decision-making. The aDBS system triggered stimulation when theta power exceeded either a fixed threshold set at 2 standard deviations (SD) above baseline mean (P1-P4) or an adaptive threshold ranging from 1.5 to 2.5 SD (P5-P9). The adaptive threshold was dynamically updated according to two rules: it decreased by 0.2 SD if no stimulation occurred within 30 seconds, and increased by 0.2 SD if more than three stimulations were triggered within 10 seconds. To prevent transient effects that could trigger a re-entrant stimulation loop during aDBS, a 2-s refractory period was imposed following stimulation offset.

The online readouts of the theta amplitude were obtained using periodogram power spectral density (PSD) estimate of the filtered LFPs, computed with a 1-second moving average Hanning window. Prior to formal testing, 4Cminute baseline recordings were obtained to determine each patient’s individualized mean and SD of theta power at the same sensing contact pair. Recordings were conducted under two conditions while patients remained relaxed and awake: 3 minutes with eyes open and 1 minute with eyes closed. This ensured a stable and representative baseline, accounting for natural fluctuations in theta activity.

### Localization

DBS electrode localization for postoperative confirmation utilized Lead-DBS v2 (http://www.lead-dbs.org)^61^. In brief, postoperative CT images were first linearly co-registered to preoperative MRI and normalized into ICBM 2009b NLIN asymmetric space using the SyN approach implemented in Advanced Normalization Tools. DBS electrodes were then localized using Lead-DBS and warped into the Montreal Neurological Institute (MNI) space using the PaCER algorithm after visual review and refinement of the co-registrations and normalizations. The patient cohort in Study I is the same as that in the previously published study, and their lead reconstruction has already been published^70^. The lead reconstruction for the patient cohort in Study II is shown in **Fig. S1a**.

### Offline Signal Processing

All data were analyzed offline using MATLAB R2023a with EEGLAB toolbox v2022.1 and custom-written scripts. One patient (P3) was excluded from the offline signal analysis due to inability to complete the recording during the sham stimulation session, although the behavioral assessment was completed. Two additional patients (P2, P8) received ghost markers making event identification difficult due to port connection issues, and were therefore excluded from offline signal analysis.

Time-frequency analyses were performed for cue-aligned, outcome-aligned, and stimulation-aligned epochs to capture the decision, reward/loss processing, and triggered stimulation phases, respectively. ERSPs were computed at both the subject and single-trial levels. For trial-by-trial analyses, ERSP changes were further normalized within each trial to enable comparison across subjects.

#### Preprocessing

Perioperative LFP signals were re-referenced offline using a bipolar montage to minimize volume conduction. Analyses focused on the sensing contact pair that was used for online feature detection during theta oscillatory state–adaptive stimulation of the STN. The data were high-pass filtered at 1 Hz to remove direct current (DC) offsets and notch filtered at 50 Hz and its harmonics using a Hamming-windowed FIR filter (pop_eegfiltnew, EEGLAB). The data were z-scored across time points. All recordings were visually inspected blind to experimental conditions, and epochs containing artifacts (e.g., cardiac activity) were identified and removed.

#### Time–Frequency Analysis

Time–frequency measures were computed using complex Morlet wavelet convolution (pop_newtimef, EEGLAB). For each trial, the LFP signal was transformed into the frequency domain using the fast Fourier transform (FFT), multiplied by the FFT of a family of complex Morlet wavelets, and then converted back to the time domain via inverse FFT. Each wavelet was defined as a Gaussian-windowed complex sine wave, where t denotes time, f frequency (4–40Hz, logarithmically spaced), and σ determines the wavelet’s width, corresponding to three cycles at the lowest frequency, with the number of cycles increasing linearly by 0.5 per octave with frequency to balance temporal and spectral resolution. Power was defined as the squared magnitude of the complex signal (*p(t) = real[z(t)J2 + ima9[z(t)J2*), normalized by conversion to decibel units. Epochs were baseline corrected for each frequency by the average power from –300 to 0ms before the cue onset. Power estimates were then obtained by averaging event-related spectral perturbation (ERSP) values within the theta band (4–8 Hz) for each trial. A sliding Wilcoxon signed-rank test (200ms window, 100ms step) was used to assess group differences, and cluster-based permutation testing was applied to correct for multiple comparisons.

#### Analysis of Stimulation Pattern

During theta state–adaptive stimulation, stimulation was delivered in real time contingent on STN theta activity. We examined whether stimulation patterns dynamically adapted to ongoing neural states during the risk-taking task, reflecting moment-to-moment changes in decision-related theta oscillations. To this end, we computed the % trials with stimulation onset using 200ms windows from 1 s before to 3 s after the decision phase and from 1 s before to 2 s after the outcome (reward/loss) and response phases, and averaged the values across subjects.

#### Determination of Theta Bursts

For single-trial analyses during the cue-elicited 3-second decision phase, theta bursts were identified following the beta burst detection framework established by Tinkhauser G et al^48^. To do this, we first extracted the time-evolved wavelet amplitude of the theta band during the cue-elicited 3-second decision phase. The burst detection threshold was defined as the 75^th^ percentile of the theta amplitude distribution. As the data has been normalized to the baseline on a trial-by-trial basis, the percentile defined threshold was applied in the trial level to enable trial-by-trial analysis. A burst was detected when instantaneous power exceeded the threshold for ≥0.1 s. Three metrics were defined and compared across stimulation conditions. Burst duration was defined as the continuous period during which the amplitude exceeded the threshold. Burst amplitude was defined as the area under the curve between the amplitude trace and the threshold. Burst density was defined as the number of bursts per second (i.e., burst count in trial-by-trial analyses).

### Hierarchical Drift Diffusion Modeling

The drift diffusion model (DDM) describes two-choice decisions as a noisy accumulation of evidence toward one of two boundaries, with parameters for drift rate (v, evidence accumulation speed), threshold (a, evidence required), non-decision time (t, pre-decision processes), and starting bias (z, initial choice bias), jointly explaining choices and reaction times^68,69^. The DDM parameter estimation is formulated as:

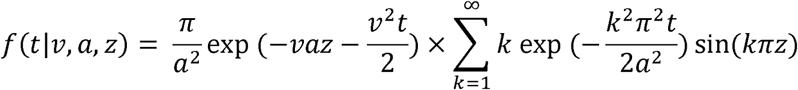

Hierarchical Bayesian estimation is applied to DDM by modeling individual parameters as drawn from group-level priors, enabling robust inference of condition-, subject-, and group-level effects using Markov chain Monte Carlo (MCMC) sampling. Parameter estimation was performed using HDDM 0.9.7 in Python 3.6, generating 11,000 samples with the first 1,000 discarded as burn-in.

We employed HDDM to examine how DBS influences decision-making under risk and uncertainty and to assess its effects on model parameters. Categorical models for different DBS conditions were fitted with trials divided by uncertainty (high vs. low) or risk (high vs. low), with decisional boundaries defined as betting versus no-betting choices. Statistical significance was evaluated using a Bayesian paired-sample t-test or repeated-measure ANOVA in JASP where appropriate, and evidence strength was quantified using Bayes factors (BF□□)^42^.

### Statistical Analysis

Reaction times (RTs) were z-scored within each subject and condition, and outliers exceeding ± 2.5 SD from the individual mean were excluded. Betting behavior was expressed as the percentage of trials in which a bet was placed, calculated as (number of bets / total trials) × 100%. Subjects without paired data and outliers exceeding ±2 SD of within-subject differences were excluded.

In study I (subacute stimulation study), behavioral scores were analyzed using repeated-measures ANOVA. One-way ANOVAs were conducted with stimulation frequency (5 Hz, 130 Hz, sham) as a within-subject factor, and two-way ANOVAs included condition (high vs. low risk or high vs. low uncertainty) and stimulation frequency as factors. Mauchly’s test was applied to evaluate sphericity, and Greenhouse–Geisser corrections were used when assumptions were violated. Post hoc comparisons were performed using protected Fisher’s least significant difference (LSD) tests. In Study II, Bets (%) and RT (s) were compared between theta oscillatory state–adaptive and sham stimulation within each condition (uncertainty: low and high; risk: low and high) and globally using paired t-tests.

Data normality was assessed using the Shapiro–Wilk test. Correlations were examined with Spearman (non-parametric) and Pearson (parametric) methods, and group differences were evaluated using the paired t test or Wilcoxon signed-rank test where applicable. Effect sizes were reported as Cohen’s d for parametric comparisons, rank-biserial correlation (rrb) for non-parametric tests, and Fisher’s z for correlation analyses.

Statistical significance was set at a 2-tailed 5% level. Data analyses were performed using SPSS 26.0, Python 3.11, or Matlab R2025a. The figures were generated using R version 4.3.1.

## Data availability

The data supporting the results of this study are available upon reasonable request from the corresponding authors.

## Code availability

The code producing the results of the study available upon reasonable request from the corresponding authors.

## Acknowledgements

This study is funded by the STI 2030-Major Projects (No. 2021ZD0200407 [to VV]), the National Natural Science Foundation of China (Grant No. 82271515 [to BS], 81971294 [to DL] and T2250710686 [to VV]), the Science and Technology Commission of Shanghai Municipality (Grant No. 20410712000 [to DL]), Medical Research Council Senior Clinical Fellowship (Grant No. MR/W020408/1 [to VV]), the SJTU Trans-med Awards Research (Grant No. 2019015 [to BS]), the Scientific and technological innovation action plan of Shanghai (Grant No. KY20211478 [to BS]), the Shanghai Municipal Science and Technology Major Project (Grant No. 2021SHZDZX [to BS]), and the Nursing Development Program of Shanghai Jiao Tong University School of Medicine (Grant No. SJTUHLXK2022 [to XQ]).

## Author contributions

DL, LW and VV initiated this work. DL and VV supervised the study. LW and VV drafted the manuscript. LW, ZZ, PH and JL collected data for the study, VV, LW, and DL designed the study. LW and VV conducted the statistical analysis and verified the data reported in the manuscript. LW, LM and VV designed the behavioral task. LW and LM set up the signal recording and close loop stimulation platform. All authors contributed to finalizing the manuscript and reviewed the final version. LW, YP, DL and VV are corresponding authors.

