## Supplementary information for "Theta Oscillatory State-Adaptive Subthalamic Stimulation Modulates Decision-Making under Risk and Uncertainty in Parkinson’s Disease"

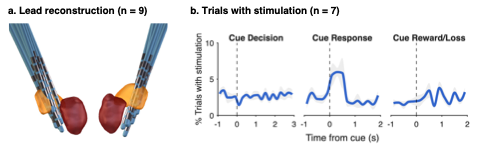

**Figure S1 Electrode localization and stimulation pattern, and beta-band dynamics. a.** Lead reconstruction across participants (n = 9) showing contact locations within the STN. **b.** Percentage of trials in which stimulation was delivered, aligned to cue, response, and outcome across different time windows (200ms, n = 7).

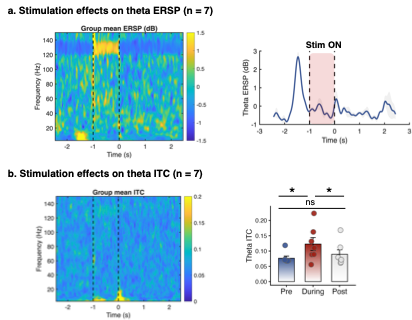

**Figure S2 Beta dynamics during decision phase under theta oscillatory state-adaptive subthalamic stimulation. a.** Left: Cue-aligned beta ERSP for high vs. low uncertainty and risk conditions during the decision phase (n = 6, subject level), showing unchanged beta-band STN activity under theta oscillatory state-adaptive subthalamic stimulation compared to sham. Paired-test p-values across the 3-s decision phase are indicated on the plot. Right: Cue-aligned beta ERSP during the decision phase (n = 6, trial level), showed beta-band STN activity was unchanged under theta oscillatory state-adaptive subthalamic stimulation compared to sham. **b.** Beta bursts (AUC, amplitude, and duration) during the decision phase (n = 6, subject level) were unchanged under theta oscillatory state-adaptive subthalamic stimulation relative to sham. **Abbreviations:** ERSP, event-related spectral perturbation; Nostim, no stimulation; CLstim, close loop stimulation; AUC, Area Under the Curve; LR, low risk; HR, high risk; LU, low uncertainty, HU, high uncertainty.

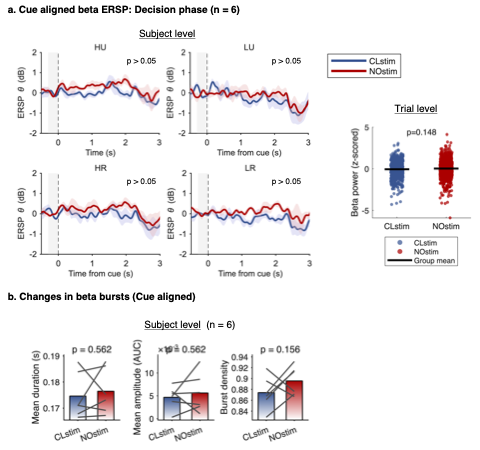

**Figure S3 Stimulation effects of Theta oscillatory state-adaptive subthalamic stimulation. a.** Time frequency plot (left) of stimulation-aligned ITC activity. Theta-band ITC (right, n = 7) aligned to stimulation onset, comparing pre-stimulation, during stimulation, and post-stimulation periods. Asterisks indicate significant differences; ns indicates non-significance. **b.** Time frequency plot (left) of stimulation-aligned ERSP. Theta-band ERSP (n = 7) aligned to stimulation onset. **Abbreviations:** ERSP, event-related spectral perturbation; ITC, inter-trial coherence; stim, stimulation.

**Table S1 Demographic information of study I**

**Notes:** NA, not assigned.

**Abbreviations:** Subj, subject; LEED, L-dopa equivalent daily dose; DBS, deep brain stimulation; DA, dopamine agonist; BDI, Beck Depression Inventory; UPDRS, Unified Parkinson’s Disease Rating Scale; UPPS-P, UPPS-P Impulsive Behavior Scale.

**Table S2 Demographic information of study II**

**Notes:** NA, not assigned.

**Abbreviations:** Subj, subject; LEED, L-dopa equivalent daily dose; DA, dopamine agonist; BDI, Beck Depression Inventory; UPDRS, Unified Parkinson’s Disease Rating Scale.
